# Identification of Novel Genes Associated with Atrial Fibrillation and Development of Atrial Fibrillation Predictive Models by Incorporating Polygenic Risk Scores and PheWAS-Derived Risk Factors

**DOI:** 10.1101/2023.08.14.23294097

**Authors:** Shih-Yin Chen, Yu-Chia Chen, Ting-Yuan Liu, Kuan-Cheng Chang, Shih-Sheng Chang, Ning Wu, Donald Lee Wu, Rylee Kay Dunlap, Chia-Jung Chan, Fuu-Jen Tsai

## Abstract

**BACKGROUND:** Atrial fibrillation (AF) is the most common atrial arrhythmia and is subcategorized into numerous clinical phenotypes. Previous studies demonstrated that early-onset AF was associated with genetic loci among the certain populations.

**OBJECTIVES:** The objective of this study was to develop AF predictive models using AF-associated single-nucleotide polymorphisms (SNPs) selected from the Genome-Wide Association Study (GWAS) of a large cohort of Taiwanese and explore whether the models posed the prediction power for AF.

**METHODS:** 75,121 total subjects including 5,694 AF patients and 69,427 normal controls with the GWAS data were included in this study. The polygenic risk scores based on AF-associated SNPs were determined and then integrated with Phenome-wide association study (PheWAS)-derived risk factors including clinical and demographic variables. The robust AF predictive models were developed through advanced statistical and machine learning techniques and then were evaluated in terms of discrimination, calibration, and clinical utility.

**RESULTS:** The results demonstrated that the top 30 significant SNPs associated with AF were located on chromosomes 10 and 16, which involved *NEURL1*, *SH3PXD2A*, *INA*, *NT5C2*, *STN1*, and *ZFHX3* genes with *INA*, *NT5C2*, and *STN1* being new discoveries in association with AF. The GWAS predictive power for AF had an area under the curve (AUC) of 0.626 (*P* < 0.001) and 0.851 (*P* < 0.001) before and after adjusting with age and gender, respectively. The results of PheWAS analysis showed that the top 10 diseases associated with discovered genes were all circulatory system diseases. The results of this study suggested that AF could be predicted by genetic information alone with moderate accuracy. The GWAS could be a robust and useful tool for detecting polygenic diseases by capturing the cumulative effects and genetic interactions of moderately associated but statistically significant SNPs.

**CONCLUSIONS:** By integrating genetic and phenotypic data, the accuracy and clinical relevance of predictive models for AF were improved. The results of this study might improve AF risk classification, enable personalized interventions, and ultimately reduce the burden of AF-related morbidity and mortality.

Graphic Abstract

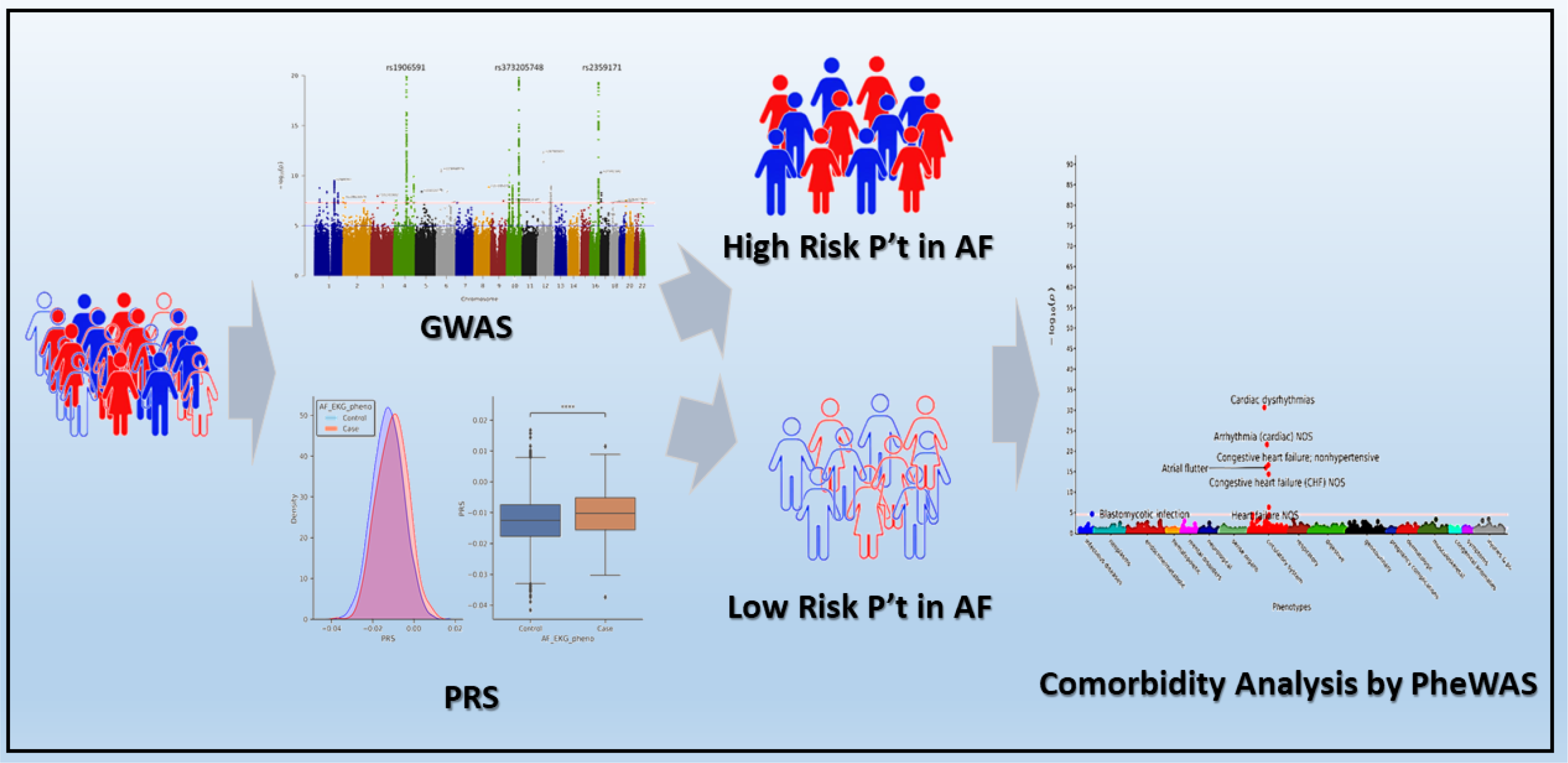

## Clinical Perspective What Is New?

- Atrial fibrillation (AF) is thought to have a heritable component, but large studies, especially Asian populations, are lacking.
- Three novel loci were identified high associations to AF in this study.
- The results of this study suggested that AF could be predicted by genetic information alone with moderate accuracy.
- The GWAS could be a robust and useful tool for detecting polygenic diseases by capturing the cumulative effects and genetic interactions of moderately associated but statistically significant SNPs.

## What Are the Clinical Implications?

- By integrating genetic and phenotypic data, the accuracy and clinical relevance of predictive models for AF were improved.
- The results of this study might improve AF risk classification, enable personalized interventions, and ultimately reduce the burden of AF-related morbidity and mortality.
- Additional prospective studies are needed to identify the genetic/genomic factors that contribute to these observed associations.
- The novel loci that had the high association to AF may have the potential to improve AF molecular genetical diagnosis.

Atrial fibrillation (AF) is a major cardiovascular disease affecting approximately 1.6% of the global population and is responsible for 20-25% of ischemic strokes and about 30% of heart failure. ^1^ AF features an irregular and fast heart rhythm, which can cause blood clots in the heart and increase the risks of stroke, ischemic heart attack, and even death. As a chronic degenerative heart disease, AF progresses from a paroxysmal to persistent type, and eventually becomes long-standing persistent and permanent AF.^2^ Among the AF patients, with more than 50% of AF being asymptomatic, the early-stage low burden paroxysmal AF is hard to be detected by a single electrocardiogram (ECG) examination. ^3^ However, once progressed to persistent AF, the patient’s heart rhythm control becomes more difficult than that in the paroxysmal AF stage, and the AF recurrence rate is significantly increased.^2^ The cause of AF is still unknown. Although it is common in people with hypertension, coronary artery disease, heart diseases, and other medical conditions such as hyperthyroidism, pneumonia, and chronic obstructive pulmonary disease, many people, especially athletes, may have it new onset without any known cause and symptoms. Recently, many studies demonstrated that AF, especially lone AF, is associated with an important genetic component.^4^ Some studies found that family history was an important factor associated to AF with the risk of AF increased by more than 40% if one parent or sibling in the family had AF.^5^ There is widespread sex, age, race, and ethnicity differences in AF epidemiology, risk factors, disease manifestations, clinical outcomes, and different reaction to therapies.^6^ In addition, Mendelian Randomization (MR) associations analysis also suggested that the body height was associated with cardiovascular disease traits including AF.^7^ All those evidence suggested that AF might be a genetically related heart disease.

Recent advancements in genomics and phenomics have provided valuable insights into the genetic and phenotypic risk factors associated with AF. Genome-wide association studies (GWAS) is an approach to identify genes associated with a particular disease by studying the entire genome of a particular large group of patients, searching for the variants of single nucleotide polymorphisms (SNPs) that occur at multiple locations across the genome associated with a specific phenotype.^8^ The GWAS for AF started in 2007.^9^ Since then, more than 93,000 AF cases and more than 1 million controls have undergone the GWAS, and more than 134 distinct AF-associated loci were identified.^10^ In addition to genetic factors, phenotypic data derived from phenome-wide association studies (PheWAS) can provide valuable insights into the non-genetic risk factors associated with AF. PheWAS analyze disease phenotypes and compared them to SNPs by using electronic medical record (EMR) data from EMR-linked databases to explore the relationships between a wide range of phenotypes and disease outcomes,^11^ which is complementary to GWAS with the ability to replicate and validate the findings of GWAS. By analyzing the association between various clinical and demographic variables and the presence of a particular disease, PheWAS can identify potential risk factors and comorbidities associated with the development of such disease. Therefore, integrating PheWAS-derived risk factors into the predictive models for a particular disease can further enhance the accuracy and clinical utility by capturing a broader spectrum of risk factors beyond genetics alone.^12^ However, there is no large sample size report of PheWAS for AF currently.

Integrating polygenic risk scores (PRS) has been used to predict the clinical phenotypes and outcomes of individuals. PRS captures the cumulative effect of multiple genetic variants across the genome, enabling the assessment of an individual’s genetic susceptibility to a specific trait or disease.^13^ Previous studies showed that an estimation of genetic susceptibility to AF was feasible with GWAS and could be used in the development of AF risk models.^14^ Moreover, integrating PRS and PheWAS-derived risk factors also demonstrated a promising approach to develop predictive models for AF.^15^ By leveraging genetic and phenotypic data, the predictive models for a particular disease have the potential to improve patient classification, identify high-risk individuals, and guide personalized prevention and management strategies. However, to take full advantage of the predictive power of the disease predictive models, a rigorous and comprehensive development and evaluation process through the combination of the data from GWAS, PheWAS, EMR, and PRS is essential.

Due to the unknown cause, insidious onset, severity of disease progression, poor prognosis, and lack of effective early diagnosis method, it is scientifically significant and clinically important to explore the potential AF mechanisms and develop an accurate method for AF prediction, early diagnosis, and prognosis evaluation. This study employed a well-characterized clinical EMR database of AF patients to develop AF predictive models by incorporating PRS and PheWAS-derived risk factors through the data of GWAS, PheWAS, and EMR. The results of this study would greatly benefit both scientists and physicians in AF study, prevention, diagnosis, and prognosis evaluation. Further, the results would potentially improve the AF patient’s risk classification, enable personalized interventions, and ultimately reduce the risks and damages to human health and the burden of AF-related morbidity and mortality.

## METHODS

### Data Resource and Patient Information

The GWAS data of 75,121 subjects including 5,694 patients with EKG confirmed AF and 69,427 patients with normal EKG from China Medical University Hospital (CMUH) (Taichung, Taiwan, ROC) during the time period of 1992 to 2020 were retrieved from the database of China Medical University Hospital Precision Medicine Project (Taichung, Taiwan, ROC). The clinical information of all involved patients was obtained from the electronic medical records (EMRs) of CMUH with the approval of CMUH ethics committees (Approval number: CMUH111-REC1-176, CMUH107-REC3-058, and CMUH110-REC3-005).

### SNP Array Data Quality Control

The TPMv1 customized SNP array (Thermo Fisher Scientific, Santa Clara, California, USA) developed by the Academia Sinica and Taiwan Precision Medicine Initiative (TPMI) projects (Taipei, Taiwan, ROC) was employed with a total of 714,457 SNPs being included in this study. PLINK1.9 (https://www.cog-genomics.org/plink/1.9/)^16^ was utilized for data analysis with the exclusions of SNPs and subjects based on the high rates of missingness per marker (geno 0.1 > 10%) for SNPs and per individual (mind 0.1 > 10%) for subjects. Variants with Hardy–Weinberg equilibrium (HWE) value less than 10-6 (HWE < 10^-6^) and minor allele frequency (MAF) value less than 10^-4^ (MAF < 10^-4^) were also filtered out. After such quality control process, 508,004 SNP variants and 75,121 subjects (5,694 AF and 69,427 normal control) remained in the study. Beagle 5.2 (https://faculty.washington.edu/browning/beagle/beagle.html) was applied for data imputation. The imputed data were further filtered by following the criteria of alternate allele dose less than 0.3 and genotype posterior probability less than 0.9.^17–19^ After quality control and imputation, a total of 9,607,262 SNP variants and 75,121 subjects (5,694 AF and 69,427 normal control) were used for analysis (The flowchart of this study was shown in Supplementary Materials Files S1).

### Genome-Wide Association Study (GWAS)

PLINK 1.9 was applied to generate summary statistics. For subjects who had EKG diagnosed AF recorded in EMR, logistic regression with multiple covariates including age and gender was performed, and the statistical significance was adjusted. Manhattan and quantile-quantile (QQ) plots with P values were generated by using R studio (https://posit.co/products/open-source/rstudio/).^20^

### Biological Pathway Analysis

The canonical pathway enriched by differential metabolites was analyzed by using the QIAGEN Ingenuity Pathway Analysis (IPA) suite (http://www.ingenuity.com) to identify relevant biological pathways and functions.^21^ The analysis was performed by integrating a group of different metabolites retrieved from Human Metabolome Database (HMDB), false discovery rate (FDR) value, and logarithmic fold change into IPA. Enrichment pathways of different metabolites were then generated based on the Ingenuity Pathway Knowledge Database (QIAGEN, Hilden, Germany).

### Phenome-Wide Association Studies (PheWAS)

The primary PheWAS analysis used 47 SNPs identified from the GWAS of AF patients to detect the potential associations between the SNPs and phenotypes extracted from the EMR. The calculated polygenic risk score (PRS) for AF was analyzed. Additionally, logistic regression was performed by using PLINK2 to examine the SNP association with phecode. A total of 97,735,180 International Statistical Classification of Diseases and Related Health Problems (ICD) version 9 (ICD-9) or version 10 (ICD-10) diagnosis codes were collapsed into 1,792 phecodes. The association between the PRS and each phecode was explored by using logistic regression models and the “PheWAS” R package.^22^ The PheWAS results were combined in a meta-analysis of multiple populations with the significance determined by using Bonferroni correction.

### AF predictive models

The regression models were then adjusted by patient’s gender, age (at the enrollment), age squared, and the top 20 principal components. Ancestry-specific PheWAS was performed in all groups, and the summarized data were then put through the meta-analysis by using an inverse-variance weighted fixed-effects model implemented in the PheWAS R package.^22^ The heterogeneity was assessed by using I^2^ and any results with excess heterogeneity (I^2^ > 40%) were excluded. The association between SNP and phecodes was also performed with false discovery rate (FDR) *P* value less than 0.01 as the significant. Thus, the thresholds for significance were set as *P* < 6.07 × 10^-5^ for critical AF variants and *P* < 4.13 × 10^-5^ for hospitalized AF variants. In this study, FDR *P* value less than 0.01 was also employed as PheWAS significant associations with increased or reduced risk for a specific condition.

### Statistical Analysis

SPSS (IBM, Armonk, New York, USA) was employed for the statistical analysis of this study.^23, 24^ The student’s t-test was used for the comparisons between two groups, while ANOVA was applied for the one-way analysis of variance among the groups. P value less than 0.05 was defined as the significant difference while P value less than 0.01 was defined as the very significant difference.

## RESULTS

### Subjects and GWAS in Atrial Fibrillation Patients

A total of 75,121 patients were included in this study with 5,694 patients who were clinically confirmed AF by EKG. GWAS was conducted in all subjects and resulted in 1,715 SNPs that satisfied with significant levels (*P* = 1×10^-5^) of AF. (the flowchart of this study was shown in Supplementary Materials File S1 and the raw data was shown in Supplementary Materials File S2 and S3). The results showed that the SNPs on chromosomes 4, 10, and 16 even reached the significant threshold (*P* < 5.0×10^-8^). By comparing the results of AF group to the controls, the top 30 significant loci were identified including *NEURL1*, *SH3PXD2A*, *INA*, *NT5C2*, *STN1*, and *ZFHX3* genes (Table 1) (raw data was shown in Supplementary Materials Files S4). The GWAS analysis results of AF were illustrated by using Manhattan plot and QQ plot (Figure 1).

**Figure 1.**
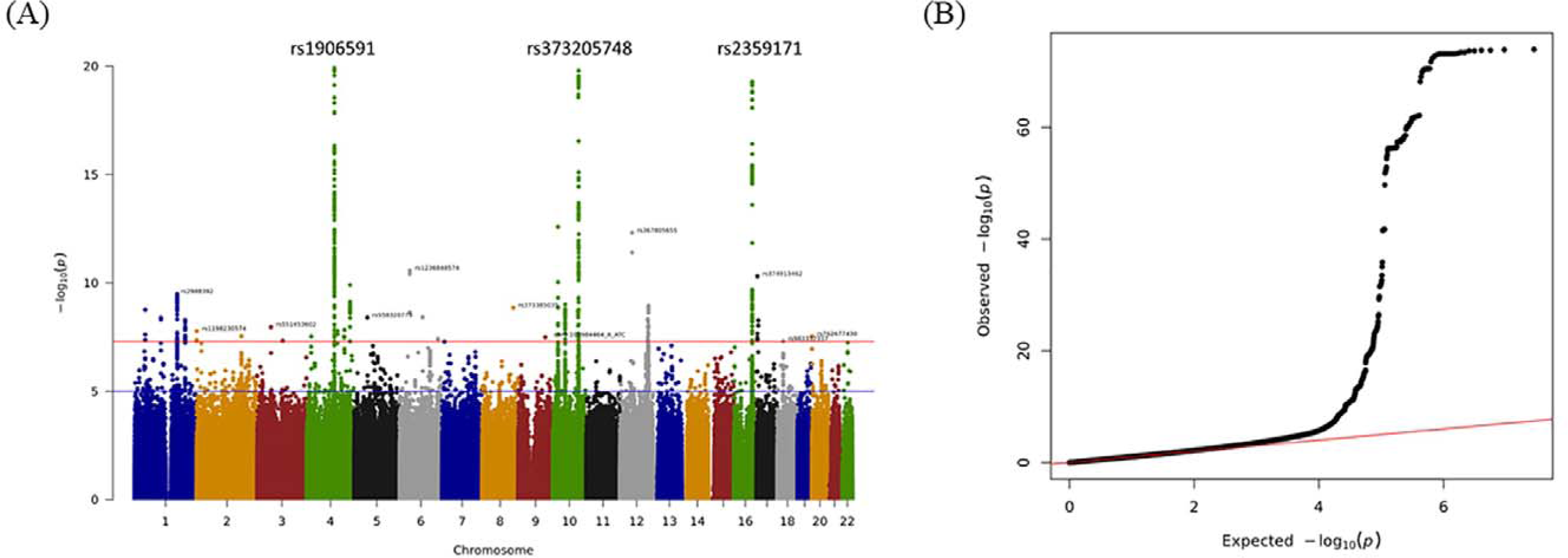
Manhattan plot (A) and quantile-quantile plot (B) for the genome-wide association study (GWAS) of ICD-10 code 148 (Ischemic Atrial Fibrillation). The upper and lower lines indicate the genome-wide significance threshold (p = 5.0 × 10^−8^) and the cut-off level for selecting SNPs (p = 1 × 10^−5^), respectively (A). The plot shows no significant deviation from the expected line, suggesting a lack of systematic biases or confounding effects in the analysis (B).

**Table 1.**
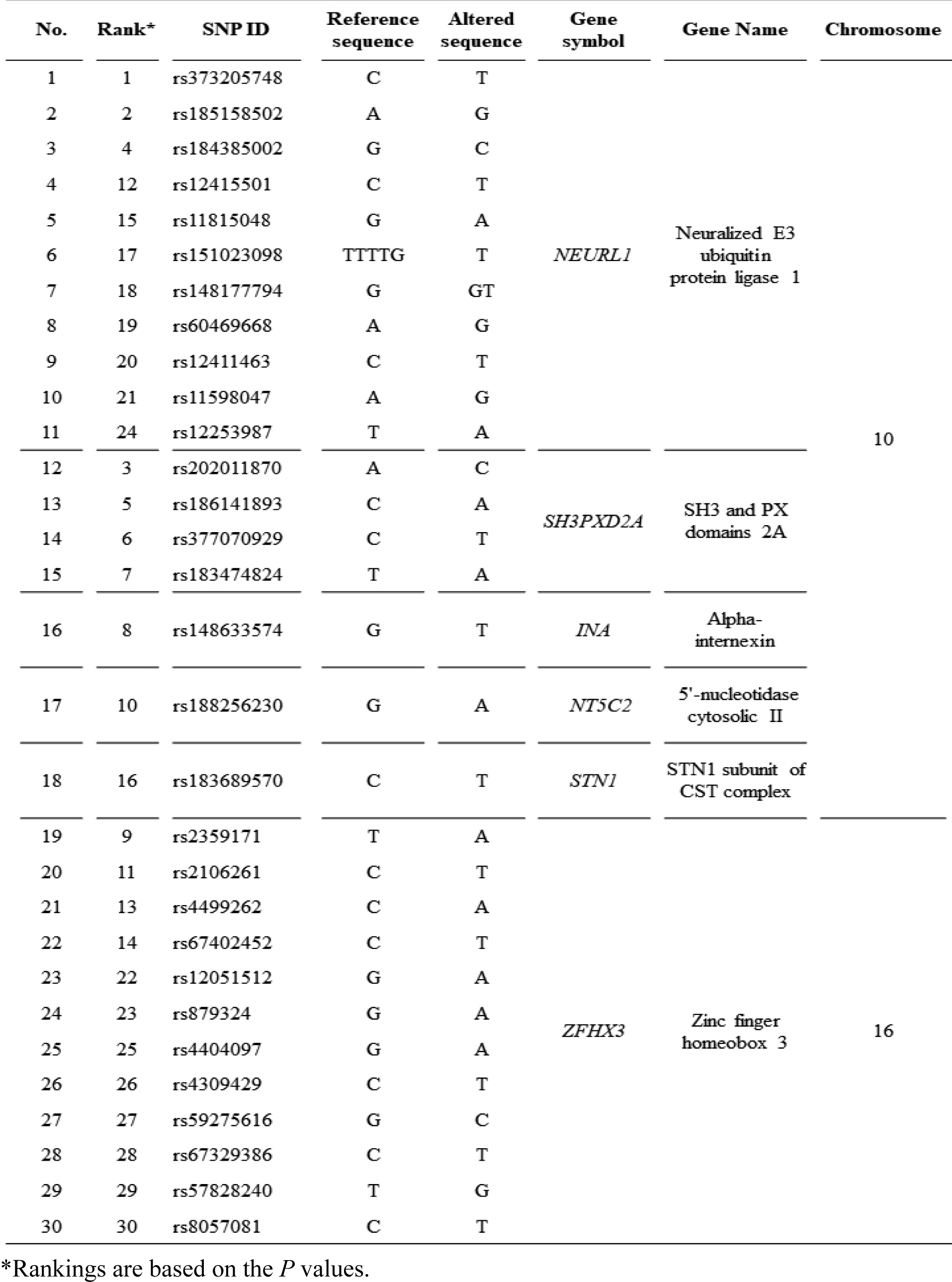
The top 30 SNPs from the discovery- and replication-GWAS.

### Identification of Polygenetic Risk Score (PRS) with Risk Predictive Models

Based on the *P* and r2 values, a set of 47 SNPs were employed to construct the optimal PRS model. Two risk prediction models for potential AF patient were established through this study with one target model (Figure 2A and 2B) and one validation model (Figure 2C and 2D). The distributions of AF PRS were demonstrated in Figures 2A and 2C with the X-axis represented PRS of AF. The PRS percentiles among AF cases versus controls were illustrated in Figure 2B and 2D with the horizontal lines, the top and bottom of each box, and the whiskers reflected the median, quartile range, and the maximum and minimum values within each group, respectively. The area under the curve (AUC) was used in this study to evaluate the performance of PRSs and the GWAS predictive power, which required individual genotypic and phenotypic data in an independent GWAS validation dataset.^25^ The GWAS predictive power for AF potential patient showed an AUC of 0.626 (*P* < 0.001) initially (Figure 3A), and after adjusted with age and gender, the GWAS predictive power for AF potential patient demonstrated an AUC of 0.851 (*P* < 0.001) (Figure 3B) (raw data was shown in Supplementary Materials Files S5).

**Figure 2.**
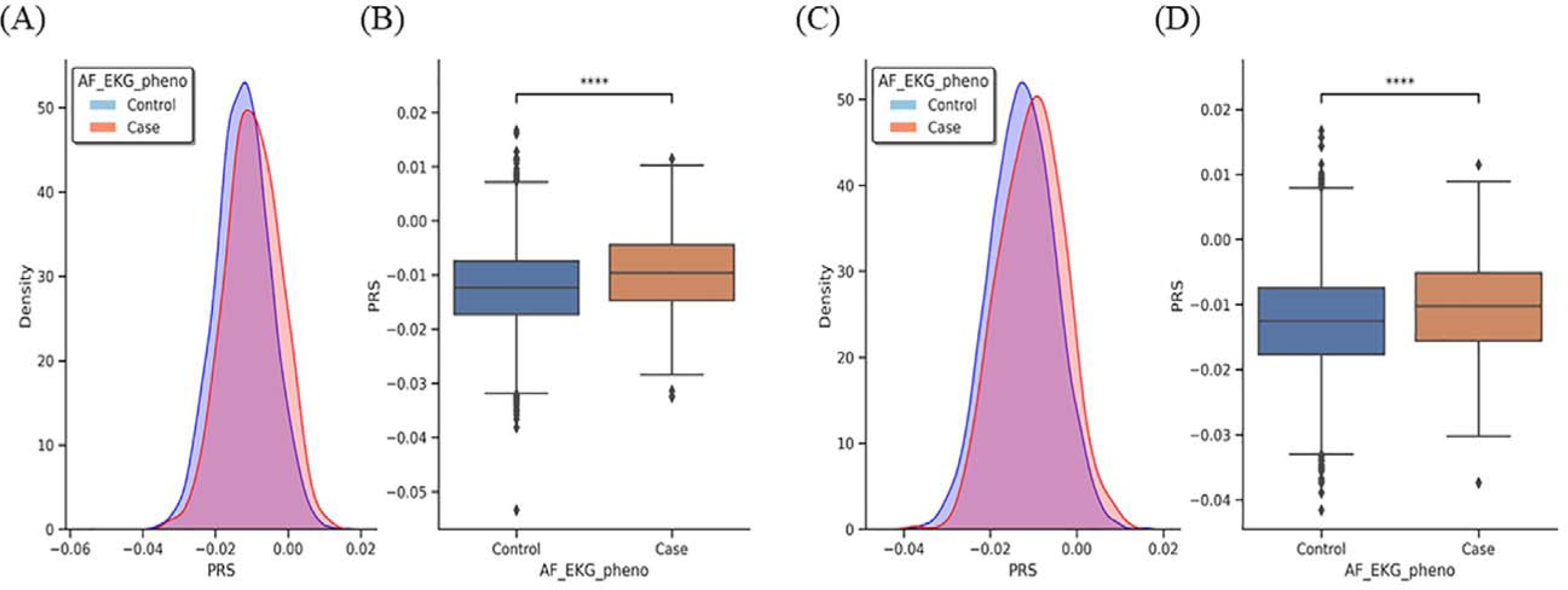
Risk analysis for atrial fibrillation (AF) based on polygenic risk score (PRS). Two AF risk predictive models were established as target model (A and B) and validation model (C and D). A and C: the distribution of AF PRS. B and D: PRS percentile among AF cases versus controls (the horizontal lines in each boxplot were the median; the top and bottom of each box were the quartile range; the whiskers were the maximum and minimum values within each group). **** represented *p*-value < 0.001.

**Figure 3.**
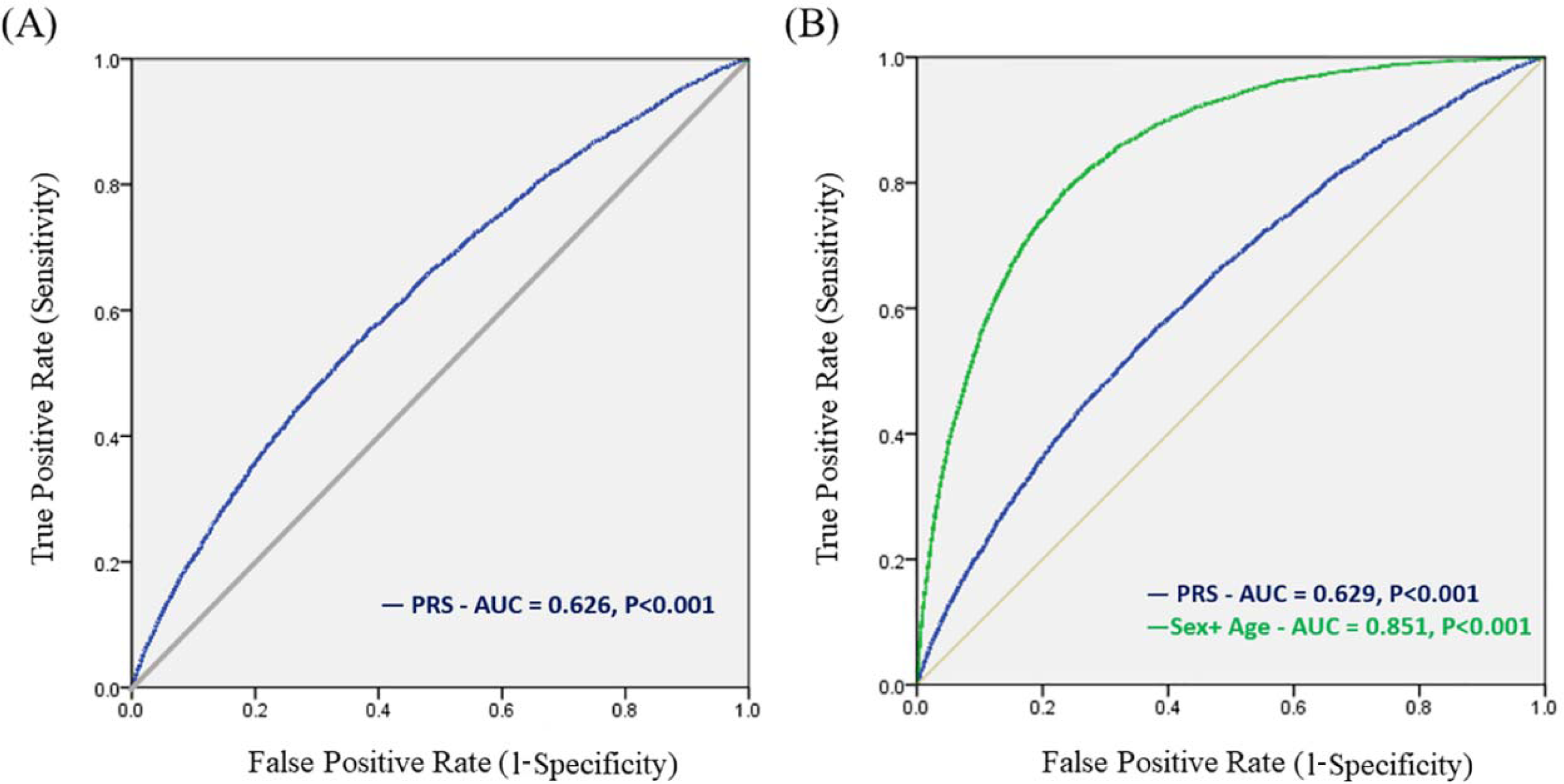
Area under the receiver-operator curves (AUC) for AF risk predictive models. (A). The GWAS predictive power for AF potential patient showed an AUC of 0.626 (P < 0.001). (B). The GWAS predictive power, after adjusted with age and gender, for AF potential patient demonstrated an AUC of 0.851 (P < 0.001).

### PheWAS in Atrial Fibrillation Patients

The association between AF PRS and 1,792 phecodes was presented by Manhattan plot. Figure 4A showed the Manhattan plot of PheWAS using top 10% PRS of AF patients *vs* the other 90% of AF patients, while Figure 4B showed the Manhattan plot of PheWAS using 4 groups of AF patients by quartile PRS. The leading top 10 diseases from the PheWAS analysis were listed in Table 2, which included atrial fibrillation and flutter, atrial fibrillation, cardiac dysrhythmias, arrhythmia (cardiac) NOS, congestive heart failure, atrial flutter, congestive heart failure (CHF) NOS, heart failure NOS, blastomycotic infection, and hypertensive heart disease (raw data was shown in Supplementary Materials Files S6 and S7).

**Figure 4.**
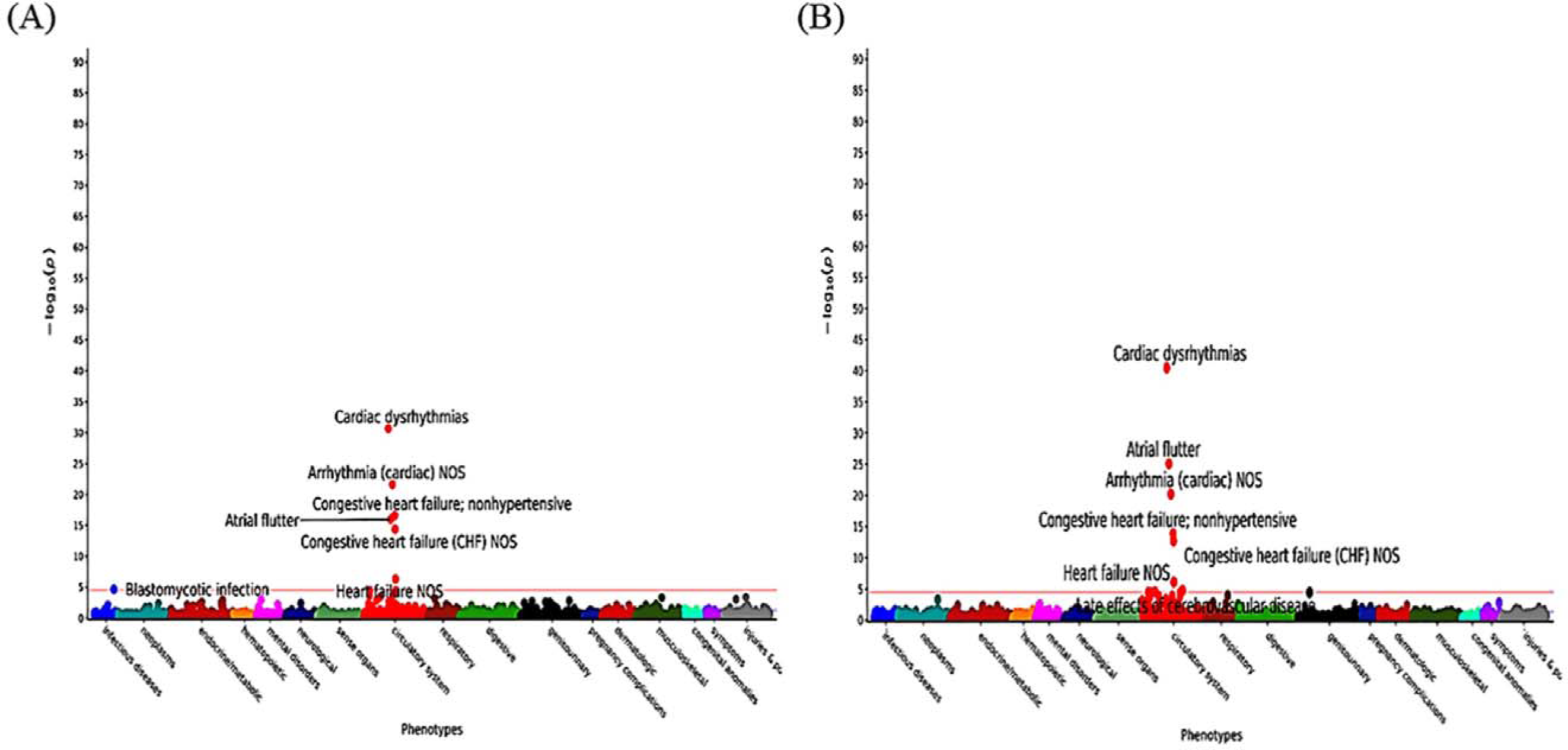
Manhattan plot of AF PRS and PheWAS presenting the association between AF PRS and 1,792 phecodes. (A): Manhattan plot of PheWAS using top 10% AF PRS patients *vs* the other 90% of AF patients. (B): Manhattan plot of PheWAS using 4 groups of AF patients by quartile PRS.

**Table 2.**
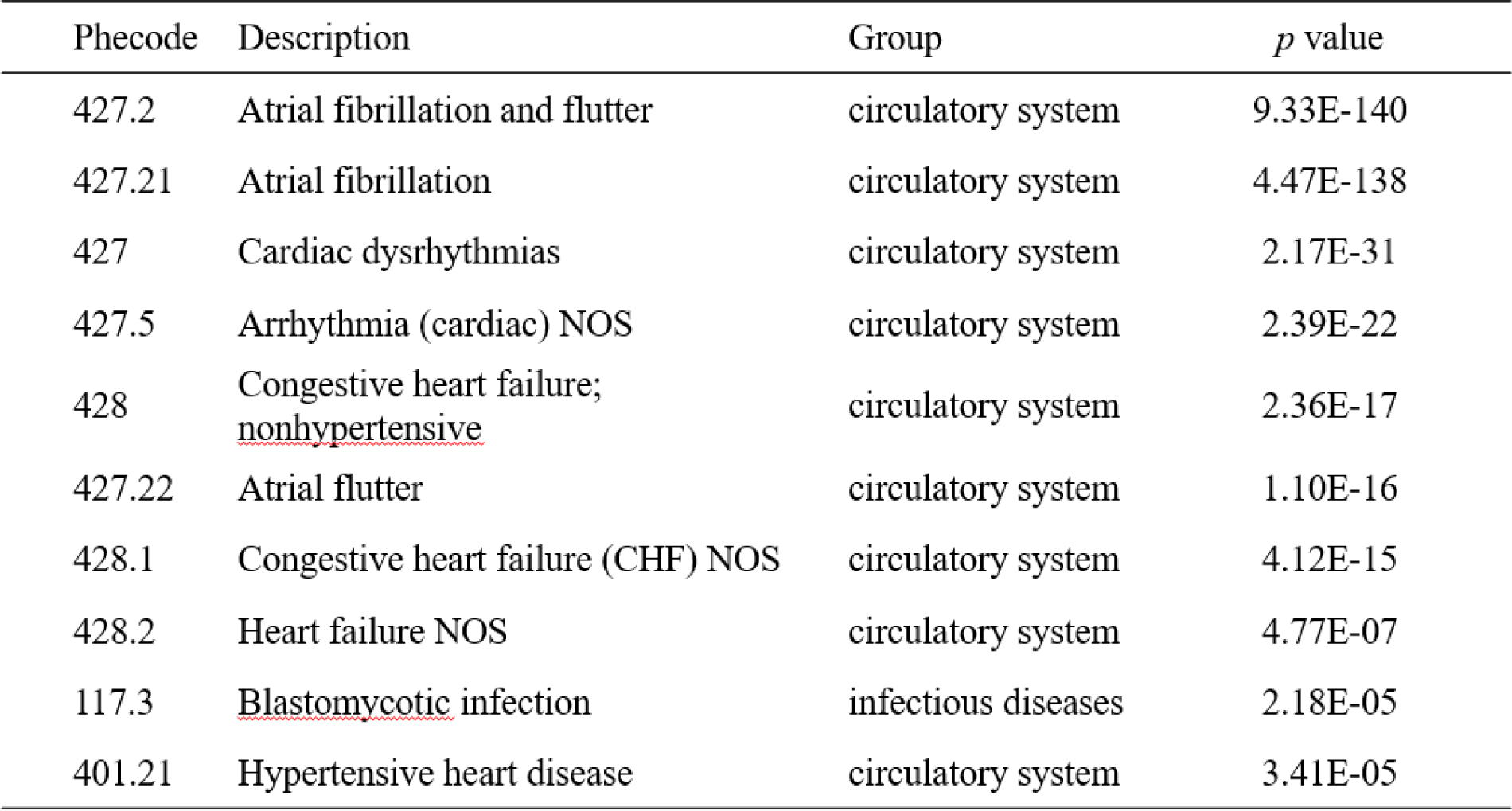
The top 10 diseases from the PheWAS analysis.

### Identification of Biological Pathways with Ingenuity Pathway Analysis (IPA)

Ingenuity pathway analysis (IPA) was employed in this study to investigate the potential biological pathways involved in the association of genetic polymorphisms and atrial fibrillation. The results demonstrated that cAMP response element-binding protein (CREB) was the major pathway associated with AF, which additionally implicated in numerous cellular pathways including gene transcription, cell growth, proliferation hypertrophy, migration, and neointimal growth (Figure 5) (raw data was shown in Supplementary Materials Files S8).

**Figure 5.**
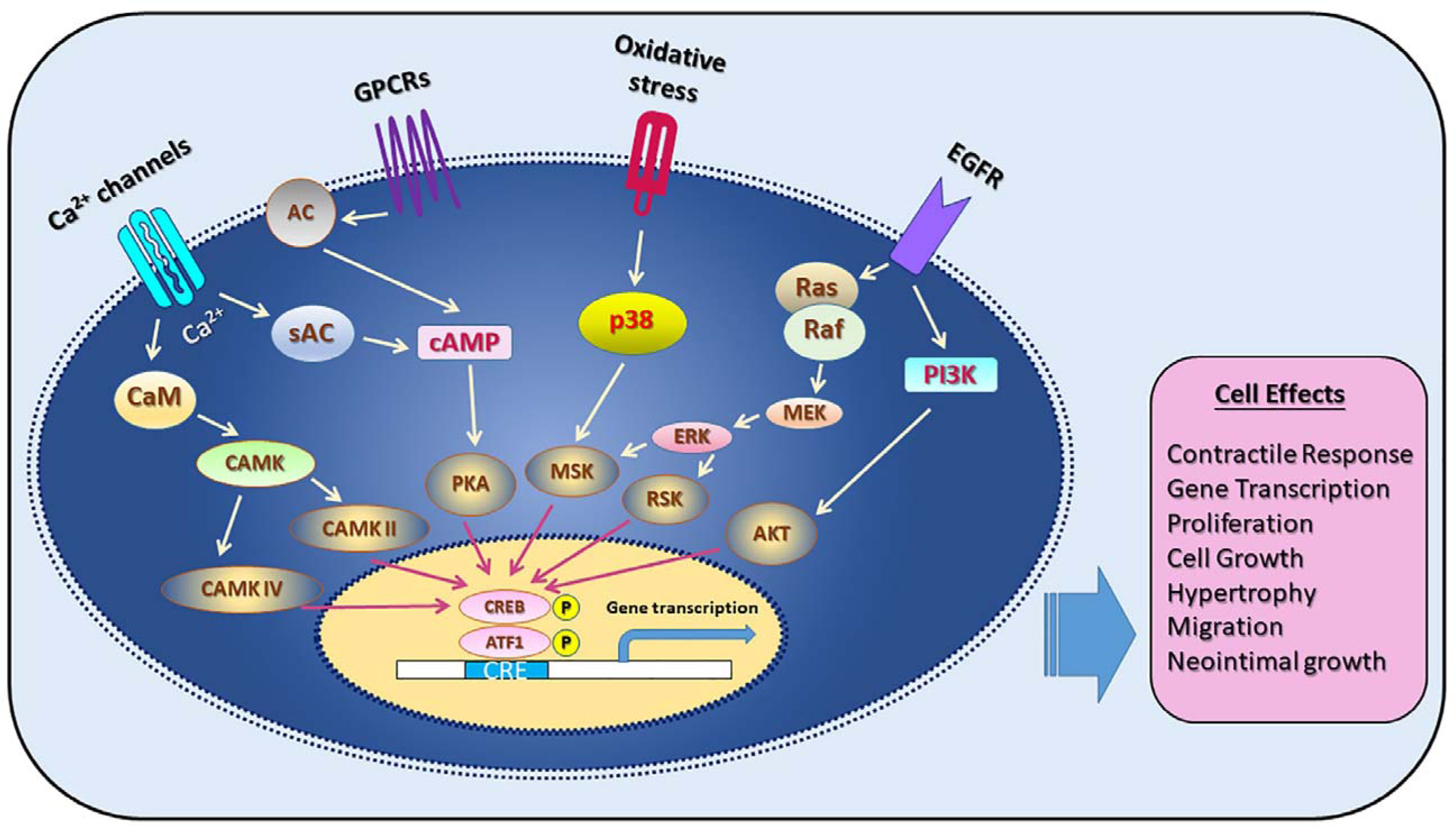
Identification and functional analysis of genes and relevant biological pathways that associated with AF by using GWAS and Ingenuity Pathway Analysis (IPA).

## DISCUSSION

### GWAS Identification of Novel Genes Associated to AF

The results of GWAS in this study identified 1,715 SNPs that associated to AF and satisfied the predetermined significant levels, with the SNPs on chromosomes 4, 10, and 16 even surpassing the stringent threshold. Among them, the genes of *NEURL1, SH3PXD2A, INA, NT5C2, STN1,* and *ZFHX3* were found among the top 30 significant loci (SNPs) with *ZFHX3* located on chromosome 16 and the others located on chromosome 10 (Table 1). According to previous studies, *NEURL1* (Neuralized E3 Ubiquitin Protein Ligase 1) showed significant allelic and genotypic associations to AF and could significantly increase genetic susceptibility to AF.^26^ *NEURL1* had been identified in AF patients of European descent and Japanese. The potential mechanism of *NEURL1* might be in prolongation of the atrial action potential duration, which had been confirmed by the knockdown of the zebrafish orthologs of *NEURL* experiment.^27^ *SH3PXD2A* (SH3 And PX Domains 2A) was reported as an AF associated gene through a GWAS study including 11,300 AF cases and 153,676 controls.^28^ Yang, et al. found that there was a replacement of Leu at 396 with Arg (L396R) in *SH3PXD2A*, which enhanced migration of macrophages and their inflammatory features, resulting in enhanced susceptibility to AF.^29^ In addition, disrupting the function of *SH3PXD2A* coding protein could affect axon guidance through locally inhibiting the degradation of the matrix in human neuron growth cone invadosome and disrupted motoneuron axons from exiting the spinal cord and extending into the periphery,^30^ which might affect the neuronal innervation of heart. *ZFHX3* (Zinc Finger Homeobox 3) has been proved an associated gene to AF.^31^ Zaw, et al. suggested that *ZFHX3* polymorphism was a risk marker for AF and AF-related phenotypes.^32^ The potential mechanism of the association between *ZFHX3* and AF was examined by knock down *ZFHX3* in atrial myocytes, and the results demonstrated the dysregulated calcium homeostasis and increased atrial arrhythmogenesis, which might contribute to the occurrence of AF.^33^

Besides the known associations of *NEURL1, SH3PXD2A,* and *ZFHX*3 to AF, this study identified three novel loci, near *INA, NT5C2,* and *STN1* genes, that also demonstrated high associations to AF. INA, also known as alpha-internexin, is a highly specific structural component of neuronal axons as a scaffolding protein in axonal and dendritic branching and growth,^34^ which plays an important role in neurite outgrowth and regulates the expression of other neurofilaments during neuronal development. However, overexpression of *INA* could induce apoptosis-like cell death,^35^ which might impact the autonomic neuronal regulation network of heart. *NT5C2* (5’-nucleotidase cytosolic II) has been found to have a significant association with coronary heart disease and hypertension.^36, 37^ Cunningham, *et al.* reported 15 significant loci associated with the risks of incident myocardial infarction or coronary artery disease, stroke, heart failure, and aortic stenosis including *NT5C2* gene.^38^ The expression of *NT5C2* is related to adenosine levels in cardiac endothelial cells and cardiomyocytes,^39^ and adenosine results in a biphasic effect on heart rate, an initial period of sinus bradycardia followed within seconds by sinus tachycardia by increasing heart cell potassium conductance,^40, 41^ which might be the potential cause of AF. *STN1* coding protein is a subunit of CST complex for telomere maintenance. The length of telomeres has been confirmed to be associated with a higher risk of ischemic heart disease both observationally and genetically.^42^ Zheng, *et al.* reported that leucocyte telomere shortening was an independent predictor of AF, and even significantly associated with recurrent AF.^43^ The study performed by Liu, *et al.* found that the leukocyte telomere length was inversely correlated with the occurrence of aging-related AF and that mitochondrial dysfunction played a role.^44^ Therefore, *STN1* could be a potential risk factor of AF, especially in aged people.

### Development of AF Risk Predictive Models

The risk predictive models of AF are useful tools for the prevention of AF episode and will help to reduce the complications and even life-threatening consequences of AF. PRS has been confirmed as a promising approach to accurately predict an individual’s risk of developing disease, which makes it possible to be employed to develop AF predictive models for the assessment of individual genetic susceptibility of AF. Two risk prediction models including the target model and the validation model were developed through this study. The distribution of PRS scores between AF patients and normal controls demonstrated significant difference, indicating the potential application values of PRS as the AF predictive markers. The results of the area under the curve (AUC) for GWAS AF predictive power showed significant results both before and after adjusting by age and gender with the AUCs as 0.626 (P < 0.001) and 0.851 (P < 0.001), respectively. Comparing the results of this study to that of previous research, the results of Wong, et al. showed that the AUCs for the maximum clumping and thresholding PRSs was 0.599 for AF after integrating age and gender into the model designing,^45^ while Torres, *et al.* reported that the AUC of age-related disease (ARD) – PRS was 0.57 for AF.^46^ The comparison results indicated that the models developed through this study showed higher AUC values than that in the past studies even in the model before adjusting by age and gender, which suggested the potential clinical applications of these models in evaluation of individuals with high risks to develop AF.

### PheWAS Analysis

PheWAS analysis of this study revealed significant associations between AF and various phenotypic traits. The Manhattan plot displayed the associations between the top 10% PRS of AF patients and other 90% of AF patients, as well as the associations between quartile PRS groups based on polygenic risk scores. Notably, the PheWAS analysis results showed that 90% (9 out of 10) of the top 10 AF PRS associated diseases were cardiovascular system diseases with the only exception of blastomycotic infection. In addition, 50% (5 out of 10) of the top 10 AF PRS associated diseases were abnormal heart rate related diseases including atrial fibrillation and flutter, atrial fibrillation, cardiac dysrhythmias, arrhythmia, and atrial flutter. These findings provided further evidence of the broad impacts of AF PRS on the health of cardiovascular system and might be the explanation of the potential comorbidities associated with the AF.^47^

### Ingenuity Pathway Analysis of Biological Pathways Associated with AF

Ingenuity Pathway Analysis (IPA) was applied to explore the biological pathways associated with AF for the purpose to identify the potential molecular signal transduction mechanisms of AF. The IPA results identified CREB pathway as the major one that associated with AF, which additionally implicated in various cellular processes including gene transcription, cell growth, proliferation, migration, and neointimal growth. Previous study suggested that AF susceptibility was associated with decreased expression of the targets of activating transcription factors (ATF)/CREB family that included a group of transcription factors related to inflammation, oxidation, and cellular stress responses.^48^ *ZFHX3*, identified as one of the 6 genes among the top 30 SNPs associated with AF in this study, was confirmed linking to CREB.^49^ In addition, the expression of *NT5C2* was related to adenosine levels in cardiac endothelial cells and cardiomyocytes,^39^ and cellular ATP production was related to the activation of CREB. Therefore, *NT5C2* and CREB together might affect the heart cell mitochondrial function^50^ and cause cardiac cell function abnormality. On the other hand, these findings also conformed the results from previous studies that AF might have a significant influence on cellular responses and played a role in modulating cell growth processes.^51, 52^ The results of this study suggested that AF associated genes might play crucial roles in regulating and controlling heart and/or neuronal cells, and potentially served as key modulators of cell growth and function. Further investigations were warranted to elucidate the precise mechanisms underlying these AF associated genes and their clinical implications in the context of AF.

### Limitations

1. Ethnic specificity: This study only included a large population from Taiwan.

There are genetic variations and differences in disease risks among different ethnicities, requiring further research to validate the accuracy and reliability of these models and newly identified loci in other populations.

1. Gene-environment interactions: This study primarily focused on the role of genes to AF, while the effects of environmental factors on those genomic changes were not considered. The onset of atrial fibrillation may result from complex interactions between genes and the environment. Therefore, future research needs to comprehensively consider environmental factors to obtain more accurate predictive models.
2. Limitations of model evaluation: Although statistical and machine learning techniques were used to develop predictive models in this study, there were still some limitations in the evaluation process. While the models’ discriminative power, calibration, and clinical utility were assessed, further studies with additional independent datasets is necessary to validate the performance of these models in real-world clinical applications.
3. Limitations in explaining genetic variations: Although the results of this study demonstrated that genetic information could be used to predict atrial fibrillation, genetic variations could only explain a portion of the disease risk. Other unknown genetic and non-genetic factors might also have significant influences on atrial fibrillation, which were not included in this study.
4. Lack of Functional Validation: Despite this study identified genetic loci associated to AF with statistical significance, further functional validation experiments were still needed to confirm the biological relevance of the identified genes and pathways and to elucidate the mechanisms through which these genetic variants contribute to AF development.
5. Limited Clinical Utility Assessment: Although the study evaluated the discrimination and calibration of the predictive models, the assessment of clinical utility, such as impact on patient outcomes or cost-effectiveness, was not investigated. Future studies should consider evaluating the real-world impact of implementing these models in clinical practice.
6. Potential biases: The data used in this study was derived from electronic medical records. Despite efforts to control confounding variables, the presence of unmeasured confounders and selection bias might not be fully ruled out which could influence the associations observed in the study and affect the validity of the predictive models.

### Conclusions

This comprehensive study integrated GWAS, PRS, PheWAS, and IPA to unravel the genetic and phenotypic aspects of AF and developed potential AF risk prediction models. The findings of this study shed lights on the genetic variants, phenotypic associations, and potential biological pathways involved in AF. The results of this study indicated the important implications for risk classification, personalized interventions, and the understanding of the underlying mechanisms of AF. Further research is needed to validate these findings and explore the clinical applications in predicting AF risks, managing potential AF patients, and improving AF patient care.

## Data Availability

The datasets used and/or analyzed during the current study available from the corresponding author on reasonable request.

## Acknowledgments

Authors would like to thank the data exploration, statistical analysis, and the support of the iHi Clinical Research Platform from the Big Data Center of CMUH, and all colleagues at the Genetic Center, Department of Medical Research, China Medical University for their feedback and technical support.

## Supplemental Material

1. S1-flowchart
2. S2-AF cohort
3. S3-AF_GWAS data
4. S4-Table 1 data
5. S5-AUC analysis
6. S6-PheWAS data-1_10PRS
7. S7-PheWAS data-2_QuartilePRS
8. S8-IPA data

